# Spatiotemporal Variations of “Triple-demic” Outbreaks of Respiratory Infections in the United States in the Post-COVID-19 Era

**DOI:** 10.1101/2023.05.23.23290387

**Authors:** Wei Luo, Qianhuang Liu, Yuxuan Zhou, Yiding Ran, Zhaoyin Liu, Weitao Hou, Sen Pei, Shengjie Lai

## Abstract

**Objectives:** The United States confronted a “triple-demic” of influenza, respiratory syncytial virus, and COVID-19 in the winter of 2022, resulting in increased respiratory infections and a higher demand for medical supplies. It is urgent to analyze each epidemic and their co-occurrence in space and time to identify hotspots and provide insights for public health strategy.

**Methods:** We used retrospective space-time scan statistics to retrospect the situation of COVID-19, influenza, and RSV in 51 US states from October 2021 to February 2022, and then applied prospective space-time scan statistics to monitor spatiotemporal variations of each individual epidemic, respectively and collectively from October 2022 to February 2023.

**Results:** Our analysis indicated that compared to the winter of 2021, COVID-19 cases decreased while influenza and RSV infections increased significantly during the winter of 2022. We revealed that a twin-demic high-risk cluster of influenza and COVID-19 but no triple-demic clusters emerged during the winter of 2021. We further identified a large high-risk cluster of triple-demic in the central US from late November, with COVID-19, influenza, and RSV having relative risks of 1.14, 1.90, and 1.59, respectively. The number of states at high risk for multiple-demic increased from 15 in October 2022 to 21 in January 2023.

**Conclusion:** Our study provides a novel spatiotemporal perspective to explore and monitor the transmission patterns of the triple epidemic, which could inform public health authorities’ resource allocation to mitigate future outbreaks.

## 1 Introduction

Since December 2019, the COVID-19 pandemic has had a catastrophic impact not only on public health but also on social and economic activities^1^. Despite the implementation of non-pharmaceutical interventions (NPIs), the availability of effective vaccines, and antiviral therapies^2, 3^, it is still a significant challenge to contain the spread of COVID-19^4, 5^. The emergence of highly transmissible variants of SAR-CoV-2^6^ and the waning protections conferred by vaccines and infection have increased the likelihood of new waves of COVID-19 outbreaks during the typical influenza season^7^. The COVD-19 restrictions over the first two years have not only suppressed the spread of influenza^8^ and RSV infections^9^, but also led to a lack of recent immune responses against them^10^. Consequently, the easing of COVID-19 restrictions might create an ideal environment for the “triple-demic” to co-circulate in the population, potentially leading to a more significant outbreak of influenza and RSV. In the winter of 2022, the Centers for Disease Control and Prevention (CDC) warned of the “triple-demic”, which refers to the simultaneous occurrence of COVID-19, influenza, and respiratory syncytial virus (RSV)^11^. Infections caused by these three respiratory diseases reached an unexpected number, occurring abnormally ahead of the typical, pre-pandemic season spike of respiratory infections^12^.

The “triple-demic” seriously challenges the public health system due to their similar transmission characteristics, clinical manifestations, and cumulative disease burden^13, 14^. For example, this “triple-demic” has overwhelmed many hospitals, leading to disrupted attendance at schools, daycares, and workplaces in the US^15^. As of October 2022, hospitalizations of infants younger than six months have reached a record high due to the emergence of RSV, with one in every 70 babies requiring hospitalization^16^. Additionally, influenza caused the highest hospital admission rates for a decade^17^. Meanwhile, COVID-19 cases have also resurged in most states, resulting in higher hospitalization rates^18^. Despite the limited time available for studying its origin, scale, and severity^19^, scholars have investigated the impact of COVID-19 and influenza co-circulation on prevalence, outcomes, and burden^20–22^. However, existing studies on the “triple-demic” are mainly clinical and overlook the general patterns of disease occurrence across space and time. It is therefore essential to analyze each epidemic and their co-occurrence to identify the most affected regions and provide valuable insights into how the “triple-demic” outbreaks relate to each other in space and time.

Space-time scan statistics (STSS) is one of the most common techniques for identifying disease clusters that characterize an outbreak, which has been widely employed in studying diseases including COVID-19^23^, inflenza^24^, dengue fever^25^ and diabetes^26^. Moreover, multivariate STSS can further identify space-time clusters of multiple diseases that occur simultaneously^27, 28^. Our study used STSS to identify spatiotemporal propagation and clusters of the emerging “triple-demic” in 51 US states. By comparing the relative risks of three infectious diseases, respectively and collectively, at the state level, our findings can contribute to understanding the scale and severity of the “triple-demic”, empowering researchers and relevant authorities to closely monitor threats to the healthcare system and formulate data-driven policies with greater precision. Ultimately, this research can facilitate better preparation and management for future public health crises.

## 2 Data and Methods

### 2.1 Data

The peak of RSV and influenza occurred unusually early in around November 2022 and a new wave of COVID-19 emerged synchronously in the US (Figure 1)^29^. We focused on the three respiratory diseases in 50 states and Washington D.C. from October 1^st^, 2022, to February 28^th^, 2023, covering the outbreaks of “triple-demic” and used the data from October 1^st^, 2021, to February 28^th^, 2022 for comparison. Given the absence of significant variations in patterns between January and February 2023, we mainly focused on the situation before February in the main text and relegated the outcomes of February to the Appendix (Table A1 & A2, Figure A1). Specifically, the COVID-19 confirmed cases, influenza cases, and diagnostic tests for RSV by testing type (i.e., PCR and Antigen) were collected from the Johns Hopkins University’s Center for Systems Science and Engineering GIS dashboard^30^, CDC FluView database^31^, and National Respiratory and Enteric Virus Surveillance System (NREVSS)^32^, respectively. The influenza case was determined by the ratio of influenza-like illness cases to all outpatient visits to sentinel physicians, multiplied by the proportion of positive respiratory viral isolates for a specific strain, weighted by the state populations^33^.

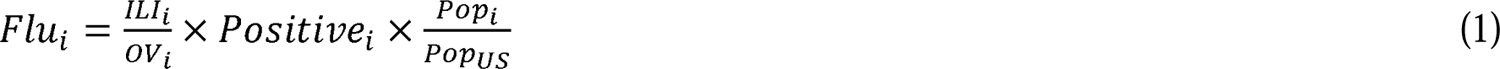

**Figure 1.**
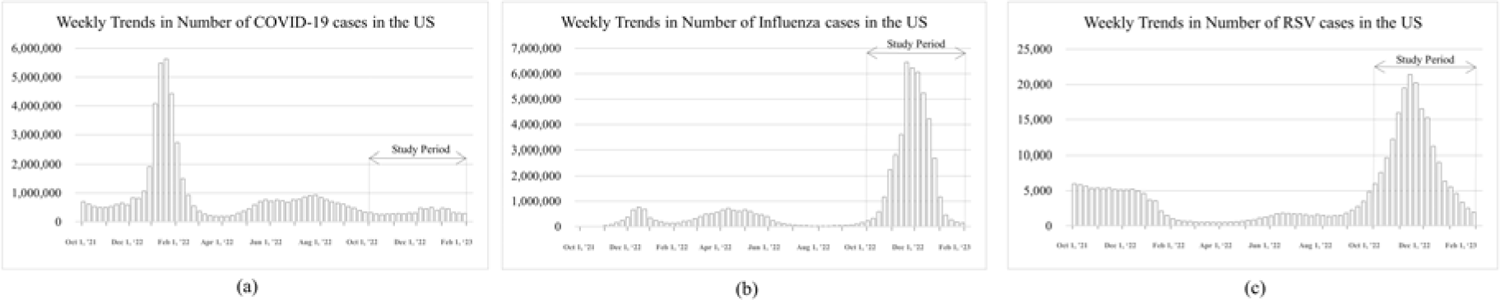
Temporal distribution of COVID-19, influenza, and RSV in the US from October 2021 to January 2023.

Where ILI_i_ is influenza-like illness cases in state i, 0V_i_ is outpatient visits to sentinel physicians in state i, Positive_i_ is proportion of positive respiratory viral isolates for a specific strain in state i, Pop_i_ is population in state i and Pop_US_ is total population in the US.

Notably, RSV data for seven states (Arkansas, Delaware, Washington D.C., New Hampshire, Rhode Island, Utah and Wyoming) were not available due to insufficient lab reporting of testing data. Moreover, we multiplied the positivity percentages and the total tests to obtain the approximate RSV cases in 44 states. Population data of 2022 in each state was obtained from the American Community Survey (ACS)^34^.

### 2.2 Space-Time Scan Statistics Method

In this study, we utilized the univariate and retrospective multivariate STSS to identify space-time clusters of COVID-19, influenza, and RSV, respectively, during the winter of 2021^35^. Subsequently, we utilized the univariate and multivariate prospective STSS to monitor the spatiotemporal progression of each epidemic and the potential triple-demic outbreak based on one-month interval during the winter of 2022^36^. We assumed that weekly cases of the three epidemics follow the Poisson distribution, proportional to the at-risk population in the United States. The null hypothesis was an inhomogeneous Poisson process with an intensity µ. The alternative hypothesis was the number of cases exceeding the expected number, which was calculated by multiplying the population of the state by the national incidence rate of the disease, as derived from the null model. We used the maximum likelihood ratio test, calculated by Equation (2), to evaluate the null and alternative hypotheses:

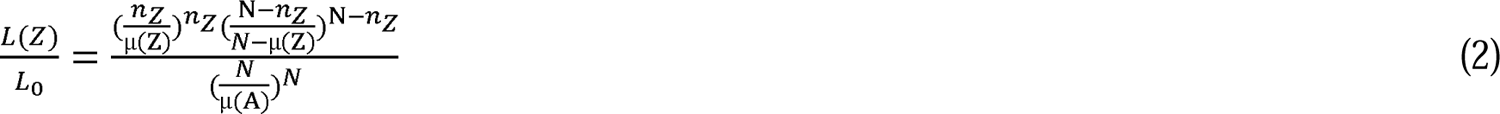

Here, L(Z) represents the likelihood function for cylinder Z, while L_O_ represents the likelihood function for the null hypotheses for cylinder Z. The likelihood ratio is calculated as the observed cases in a cylinder n_Z_ divided by the expected cases µ in a cylinder Z to the power of the observed n_Z_, multiplied by the observed cases outside the cylinder divided by the expected cases outside the cylinder. The numerator is then divided by the quotient of the total number of observed cases for the entire study area N across all time periods A to the power of the total number of observed cases. If the likelihood ratio is greater than 1, the cylinder will have an elevated risk. The space-time scan statistic uses different cylinder sizes, and the cylinder with the highest likelihood ratio (maximum) is the most likely cluster.

To identify the co-occurrence of disease outbreaks during our study period, we used the multivariate space-time Poisson scan statistic^36^. We computed the log-likelihood ratio (LLR) for each disease within each cylinder. The multivariate statistic sums the LLRs for different variables in a particular cylinder, producing a new LLR for that cylinder. Finally, the cluster with maximum summed LLR is reported as the most likely cluster, which is calculated by Equation (3):

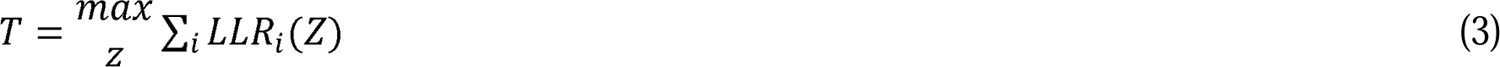

Where T is the most likely cluster within cylinder Z for a particular disease i.

In space-time analyses, two parameters were defined for cluster specification: the maximum spatial cluster size within the spatial window and the maximum temporal cluster size within the temporal window. Given the variation in population densities across the United States, the spatial cluster size was restricted to 30% of the population at risk. Spatial clusters were also defined to cover 25% and 35% of the total population at risk. However, for 35% of the total population at risk, SaTScan generated clusters that were excessively large and had relatively small relative risks (RR) when compared to those generated by using 30% of the total population at risk. On the other hand, for 25% of the total population at risk, the number of clusters was greater than that generated by using 30% of the total population at risk. The total number of observed cases and expected cases were the same for both clusters, which indicates that the maximum cluster size was sufficiently limited by 30% of the population at risk. The analyses were conducted using a maximum spatial cluster size of 30% of the population at risk in the spatial window and a maximum temporal cluster size of 50% of the study period in the temporal window. Moreover, we set a minimum of two cases in each cluster to ensure the presence of at least two cases in each cluster and employed one-week time aggregation for continuous monitoring. Monte Carlo simulations were conducted (n = 999) for each model run to evaluate the statistical significance, and clusters with a *p-value* less than 0.05 and no geographical overlap were subsequently reported as most likely and secondary clusters. Our analysis was conducted in SaTScan™^37, 38^.

Additionally, we mapped the spatiotemporal dynamics of states with multiple-demic risks based on the relative risk (RR) to illustrate the propagation of triple-demic at state level during the winter of 2022. RR is a measure of the likelihood of contracting the disease in a location compared to all other locations, which is calculated as:

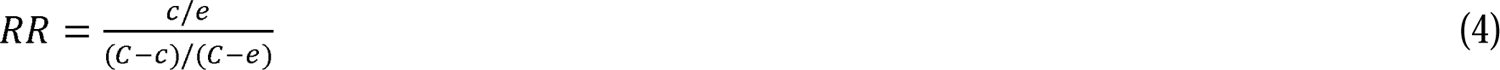

Where *c* is the total number of observed cases in a state, e is the total number of expected cases in a state, and *C* is the total number of observed cases in U.S. Specifically, RR > 1 denotes high-risk clusters with more observed cases than expected, while RR < 1 indicates low-risk clusters.

## 3 Results

### 3.1 Spatiotemporal progression of respiratory infections in the US, Oct 2021-Feb 2023

From October 2021 to February 2023, cases of COVID-19, influenza, and RSV in each state presented various dynamics over time (Figure 2). The number of COVID-19 cases experienced a dramatic growth in each state from January 2022, following which it showed a slight fluctuation from April to September 2022 and maintained at a relatively low level but not vanished during the winter of 2022. Additionally, several states in the central US have experienced potential multiple-demic risk during the winter of 2021 as the relatively high number of RSV cases and influenza cases (e.g., South Dakota, Wisconsin, Minnesota, Kansas, Idaho, etc.). Moreover, cases of influenza and RSV in many states saw noticeably sharp increases during the winter of 2023 despite the seasonal changes of the diseases, which resulted in the potential outbreak of triple-demic during the winter of 2023. The paired t-test result further showed that compared to the situation during the winter of 2021, the average number of COVID-19 cases in the whole country significantly decreased (change = −537315), while the average number of influenza cases (change = 733481) and RSV cases (change = 2534) in the whole country significantly increased during the winter of 2022.

**Figure 2.**
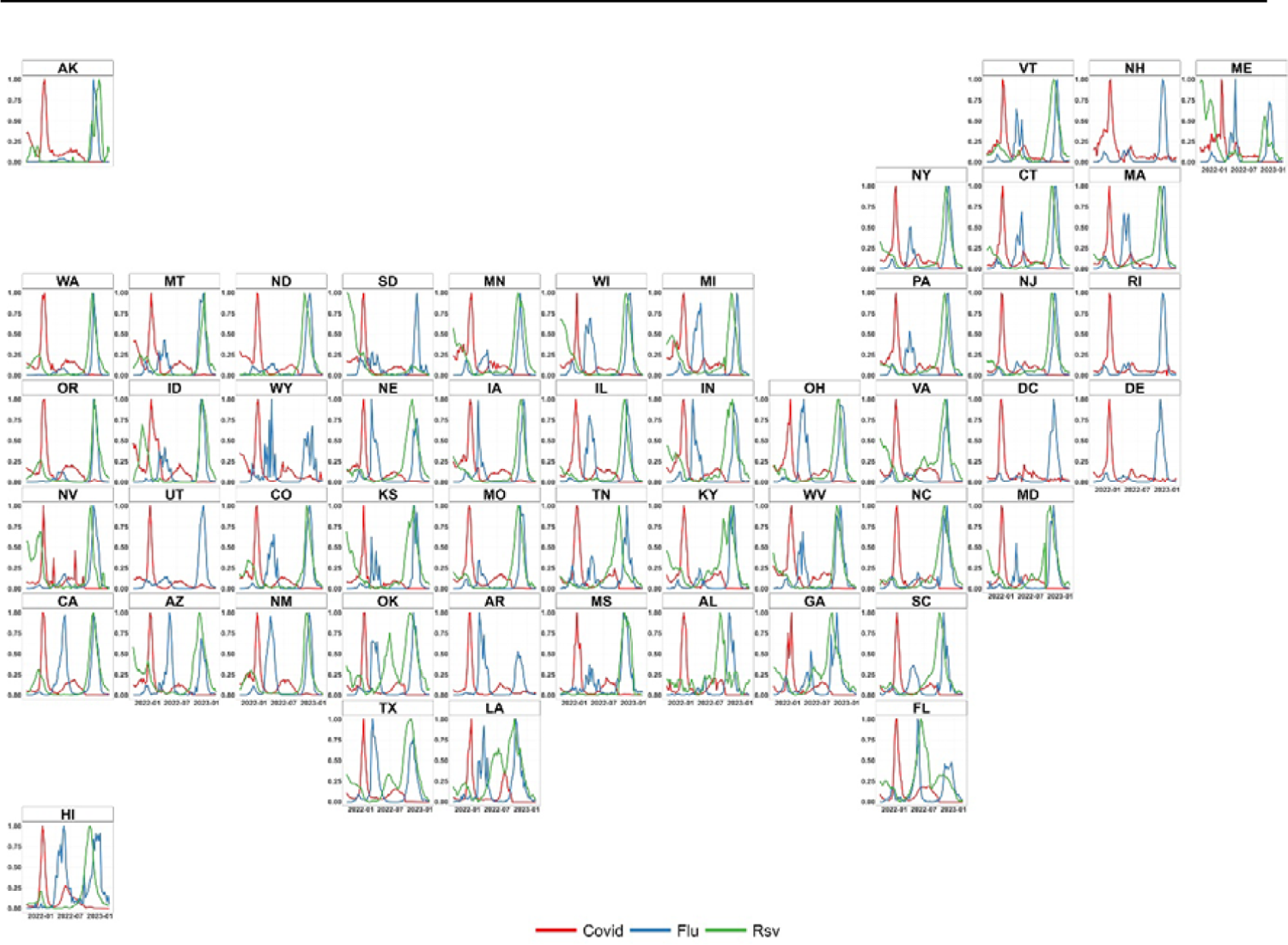
Spatiotemporal variation of respiratory infections in US from Oct-2021 to Feb-2023.

Table 2 summarizes the space-time clusters existing during the winter of 2021. The whole country experienced a severe COVID-19 situation in January 2022, with 3 high-risk clusters emerging in most states. Specifically, the most likely cluster (cluster 1), with the highest RR of 3.56, emerged in the western US, while 2 other high-risk clusters (RR=3.38, and RR=2.63) emerged in 28 eastern states (Table 2 & Figure 3a). As for influenza, a high-risk cluster (RR=3.32) emerged in 18 central and western states between November and December 2021, while an extremely low-risk cluster (RR=0.026) occurred in 16 eastern states from January to February 2022 (Table 2 & Figure 3b). Besides, 2 high risk clusters and 2 low-risk clusters of RSV emerged during the winter of 2021. Cluster 1, which had an extremely high RR of 28.32, was detected in Wisconsin from early October to early December 2021. This indicated that people living in Wisconsin were 28.32 times more likely to be infected with RSV during this period. Additionally, another high-risk cluster (cluster 3, RR=2.40) were identified in 17 western states between November and December 2021 (Table 2 & Figure 3c). However, 2 low-risk cluster (cluster 2 & 4) with an RR of 0.15 and 0.20, respectively, were identified in the eastern US from the late December 2021 to February 2022, indicating that the RSV situation mitigated in many eastern states during this period (Table 2 & Figure 3c).

**Figure 3.**
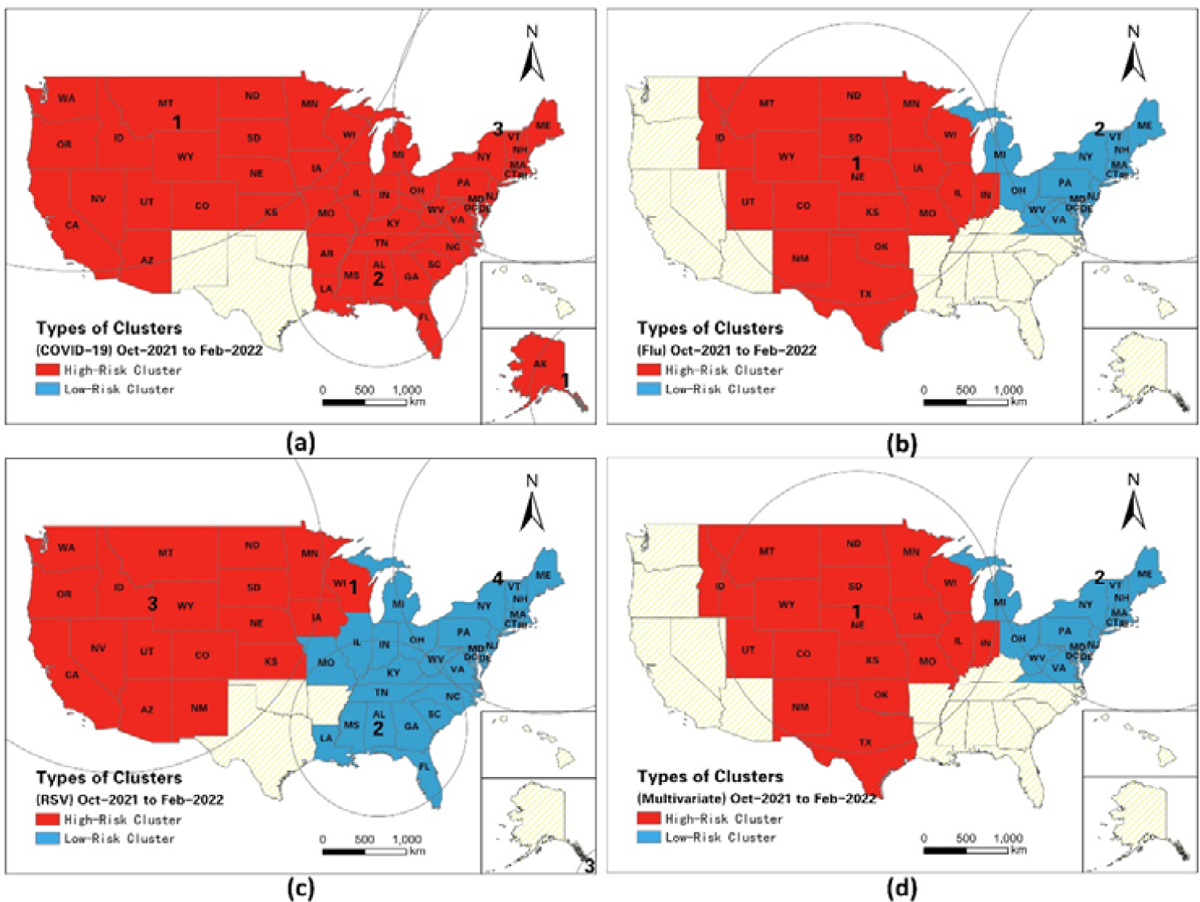
Spatiotemporal Patterns of Retrospective Space-Time Clusters during the winter of 2021.

**Table 1.** Changes in cases of each respiratory infectious disease (2021 winter vs. 2022 winter).

|  | COVID-19 (N=51) | Influenza (N=51) | RSV (N=44) |
| --- | --- | --- | --- |
| **2021 Winter (T1) [Mean (SD)]** | 673590 (103229) | 97674 (18930) | 1936 (539) |
| **2022 Winter (T2) [Mean (SD)]** | 136275 (22514) | 831155 (149851) | 4470 (944) |
| **Difference (T2-T1) [Mean (SD)]** | -537315 (84041) *** | 733481 (133862) *** | 2534 (535) *** |

Note: The paired t-tests were conducted to compare the number of cases for 2021 winter and 2022 winter. The results are labeled with asterisks in the rows showing the change: \*\*\* $p < 0.001$ .

**Table 2.**
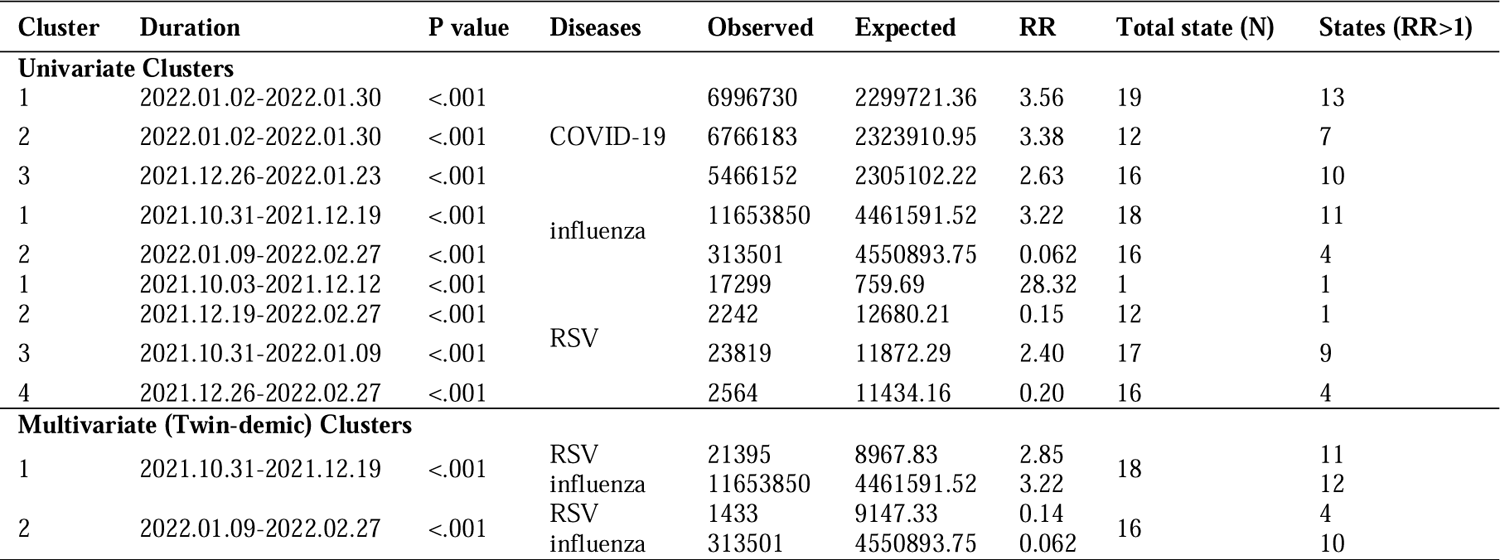
Space-time univariate clusters from October 2021 to February 2022.

For the retrospective situation of multiple demic risk during the winter of 2021, a high-risk twin-demic cluster of RSV and influenza, with an RR of 2.85 and 3.22 for RSV and influenza, respectively, occurred in 18 central and western states from November to December 2021 (Figure 3d). This implied that the population within the cluster experienced higher risk of both RSV and influenza infections. On the contrary, a low-risk twin-demic cluster of RSV and influenza (RR of RSV=0.14, RR of influenza=0.062) occurred in 16 eastern states from January to February 2022, during which the situation of COVID-19 deteriorated, indicating that the twin-demic outbreak mitigated but COVID-19 outbreak emerged during the late winter (Figure 3a & Figure 3d). In addition to twin-demic cluster of RSV and influenza, no other types of twin-demic clusters and triple-demic clusters were detected during the winter of 2021.

### 3.2 Spatiotemporal Propagation of Space-Time Clusters during the winter of 2022

#### 3.2.1 Spatiotemporal Propagation of Univariate Clusters

Figure 4 present univariate space-time clusters that emerged between October 2022 and January 2023 at a one-month interval. In October, two clusters of high COVID-19 risk, had RR of 1.77 and 1.41, respectively, were found in the northeastern US (Figure 4a). Two high-risk influenza clusters with RR of 7.28 and 3.02 emerged in nineteen southeastern states, while a large low-risk cluster emerged in the western US (Figure 4e). Besides, five dispersed high-risk RSV clusters were identified in 13 US states, with Hawaii showing the highest relative risk (RR=9.60) (Figure 4i). In November, three COVID-19 clusters (RR=1.55, RR=1.88, and RR=1.19) were identified in five southwestern states and nine eastern states (Figure 4b), and all low-risk clusters of influenza dissipated and were succeeded by 2 high-risk clusters (RR=2.56 and RR=2.62) situated in the central and eastern regions of the US (Figure 4f). The high-risk cluster of RSV that was previously confined to the northeastern US expanded to encompass 10 additional western states, with an RR change from 5.80 to 3.47 (Figure 4j). In December, the situation deteriorated significantly as high-risk clusters of the three diseases proliferated. For high-risk COVID-19 clusters, in addition to the high-risk clusters in November, a new high-risk cluster were found in Louisiana, with an RR of 5.97 (Figure 4c). High-risk influenza clusters spread to the northern region (Figure 4g), and 3 high-risk clusters emerged in Wisconsin (RR=9.31), Oregon (RR=6.94), and 3 states (Colorado, Nebraska and New Mexico) in the central US (RR=2.96) (Figure 4k). In January, most regions had low risk, but the eastern coast had persistent high-risk COVID-19 exposure (Figure 4d), and the high-risk cluster in Louisiana expanded to include Arkansas, Mississippi, and Alabama, with an RR changing to 1.67. The high-risk influenza cluster disappeared (Figure 4h), but high-risk RSV clusters remained in Wisconsin (RR=5.61) and Oregon (RR=4.67) (Figure 4i), with relief arriving by February (Table A1 & Figure A1). More details of the dynamics for univariate clusters during the winter of 2022 are show in Table A3.

**Figure 4.**
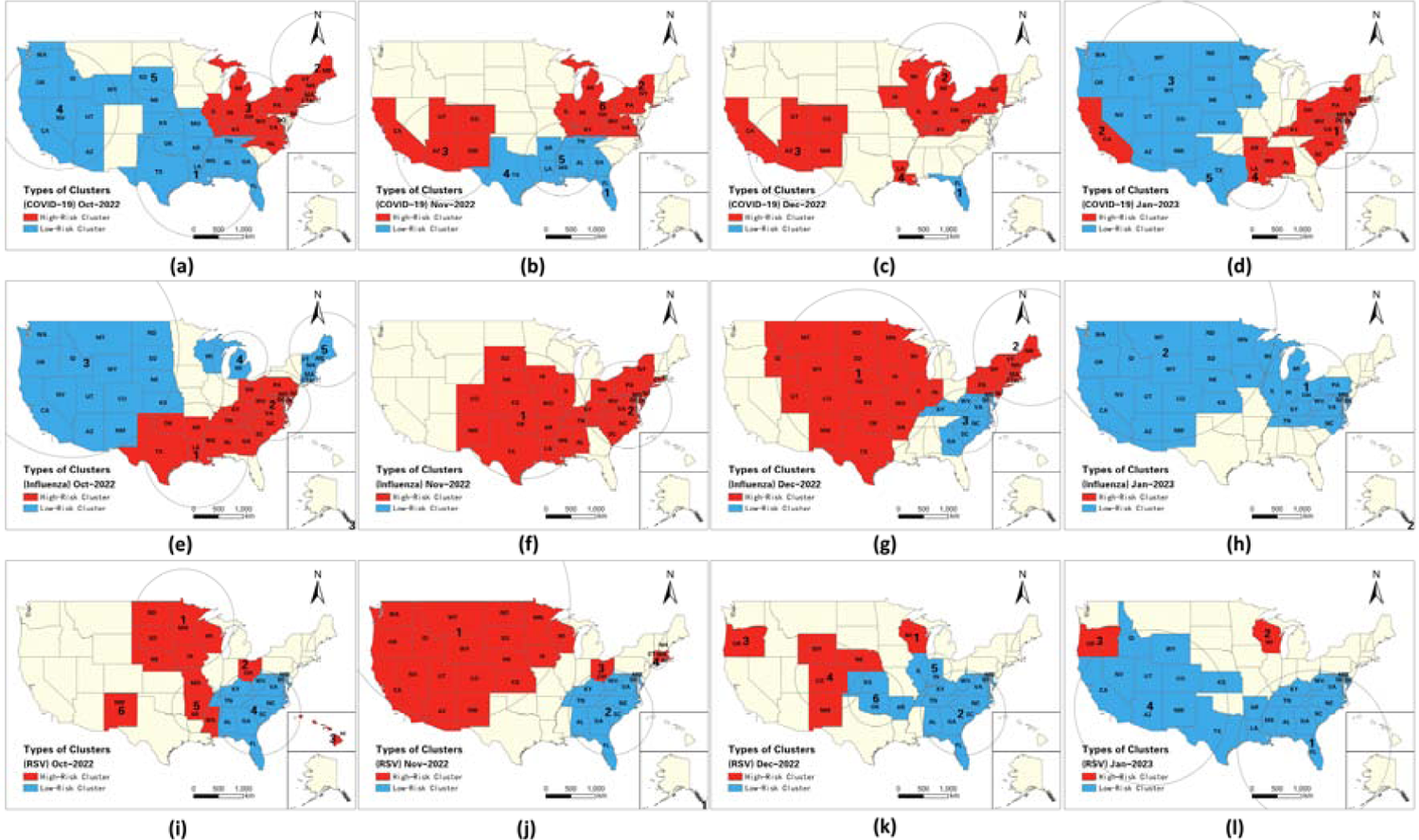
Spatiotemporal Patterns of Univariate Space-Time Clusters during the winter of 2022.

#### 3.2.2 Spatiotemporal Propagation of Multivariate Clusters

Table 3 and Figure 5 illustrate the spatiotemporal dynamics of multivariate space-time clusters. In late October, a high-risk twin-demic cluster consisting of eight southeastern states emerged (Figure 5a), exhibiting relatively high infection risks of both RSV (RR=1.05) and influenza (RR=7.28). Furthermore, a high-risk cluster of triple-demic comprising 11 eastern states emerged with an RR of 1.60 for RSV, 3.02 for influenza, and 1.41 for COVID-19, which raised concerns of triple-demic outbreaks in this region. In contrast, a large low-risk twindemic cluster of influenza and COVID-19 emerged in the western US, demonstrating relatively low risks of influenza (RR=0.58) and COVID-19 (RR=0.80) infections. In early November and late November, two high-risk twindemic clusters surfaced. The first cluster consisted of 15 states located in central and southern regions, with six states exhibiting relatively high risks (RR > 1) for RSV and seven states showing relatively high risks for influenza (Figure 5b). This cluster had an RR of 1.26 for RSV and 2.56 for influenza. The second cluster comprising 15 states in the eastern coastal regions of the US had an RR of 2.62 for influenza and 1.18 for COVID-19. From late November to December, a large high-risk triple-demic cluster (cluster 1 in Figure 5c) emerged in the central US, evolving from the previous twindemic cluster of RSV and influenza. This cluster had an RR of 1.59 for RSV, 1.90 for influenza, and 1.14 for COVID-19. Meanwhile, 9 states located in the eastern coast formed a high-risk twindemic cluster of influenza (RR=2.13) and COVID-19 (RR=1.56) (cluster 2 in Figure 5c), indicating that individuals residing in the cluster experienced a high risk of both influenza and COVID-19 infections. However, the southeastern US saw a low risk twindemic cluster of RSV (RR=0.19) and influenza (RR=0.39), suggesting that the situation of RSV and influenza had improved in these areas. Starting from early January, a low-risk twindemic cluster of RSV (RR=0.12) and COVID-19 (RR=0.10), emerged in 13 states in the eastern US (cluster 1 in Figure 5d). This finding suggests that the triple-demic situation had mitigated in these states between January 02, 2023 and the end of January. Additionally, a low-risk triple-demic cluster emerged in 18 states in the western and central regions with an RR of 0.61 for RSV, 0.10 for influenza, and 0.80 for COVID-19 (cluster 2 in Figure 5d). Continued vigilance is needed as 11 states within the low-risk triple-demic cluster still experienced a relatively high RR for influenza, another 11 states still had a relatively high RR for RSV, and 4 states still had a relatively high RR for COVID-19.

**Figure 5.**
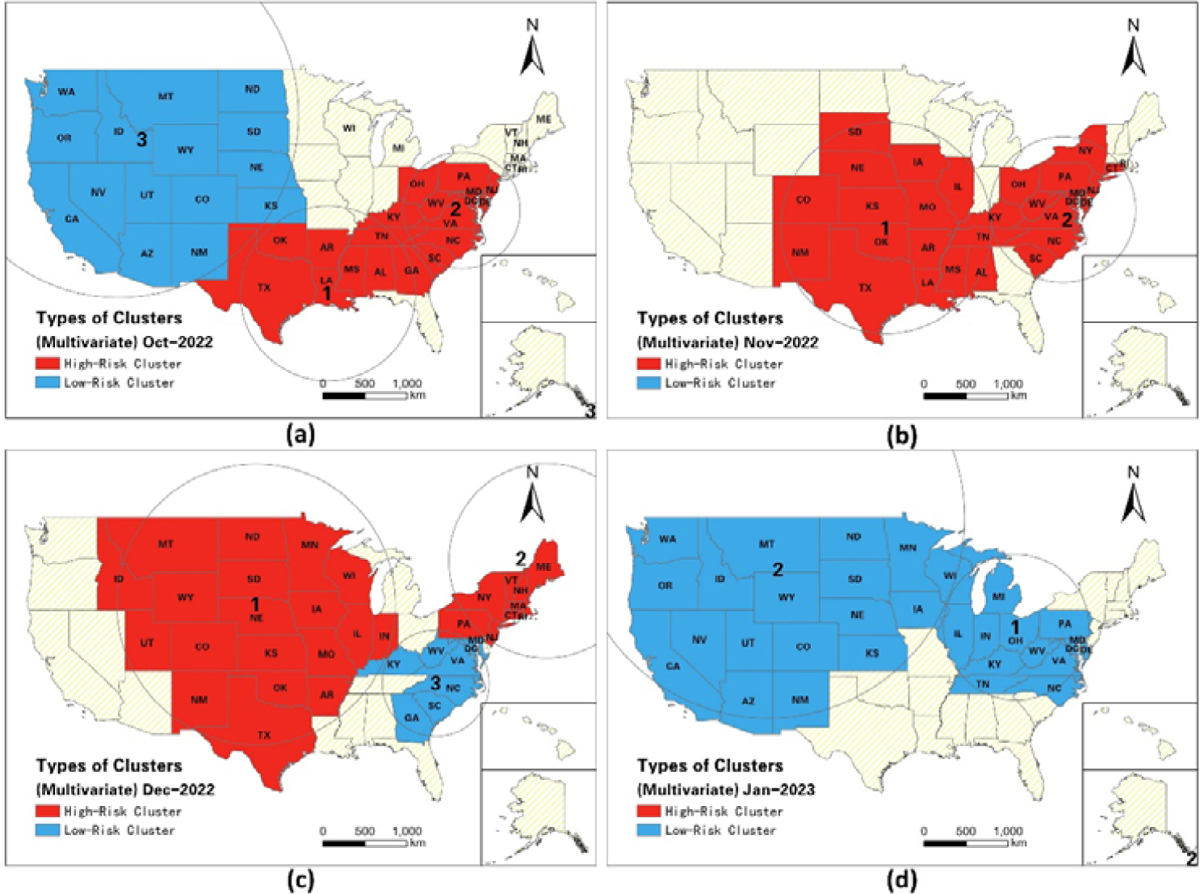
Spatiotemporal dynamics of Multivariate clusters.

**Table 3.**
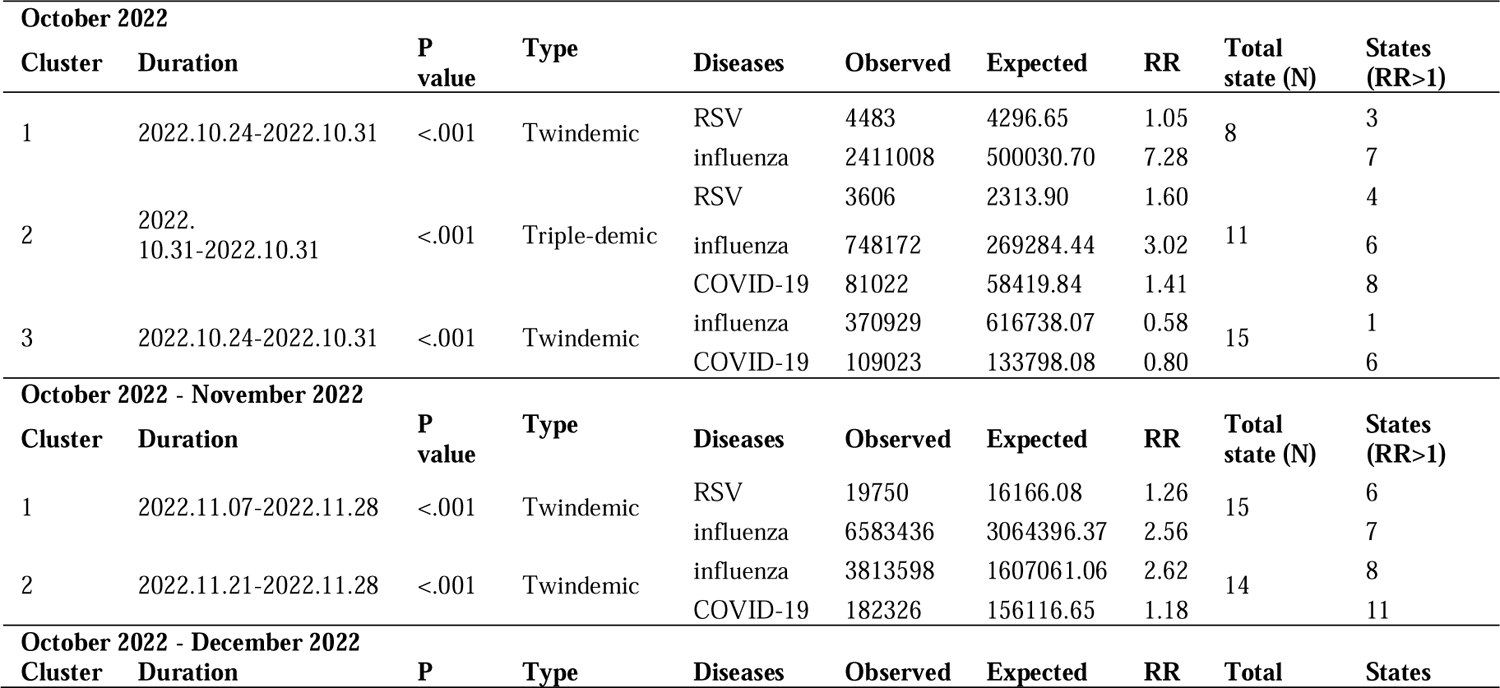
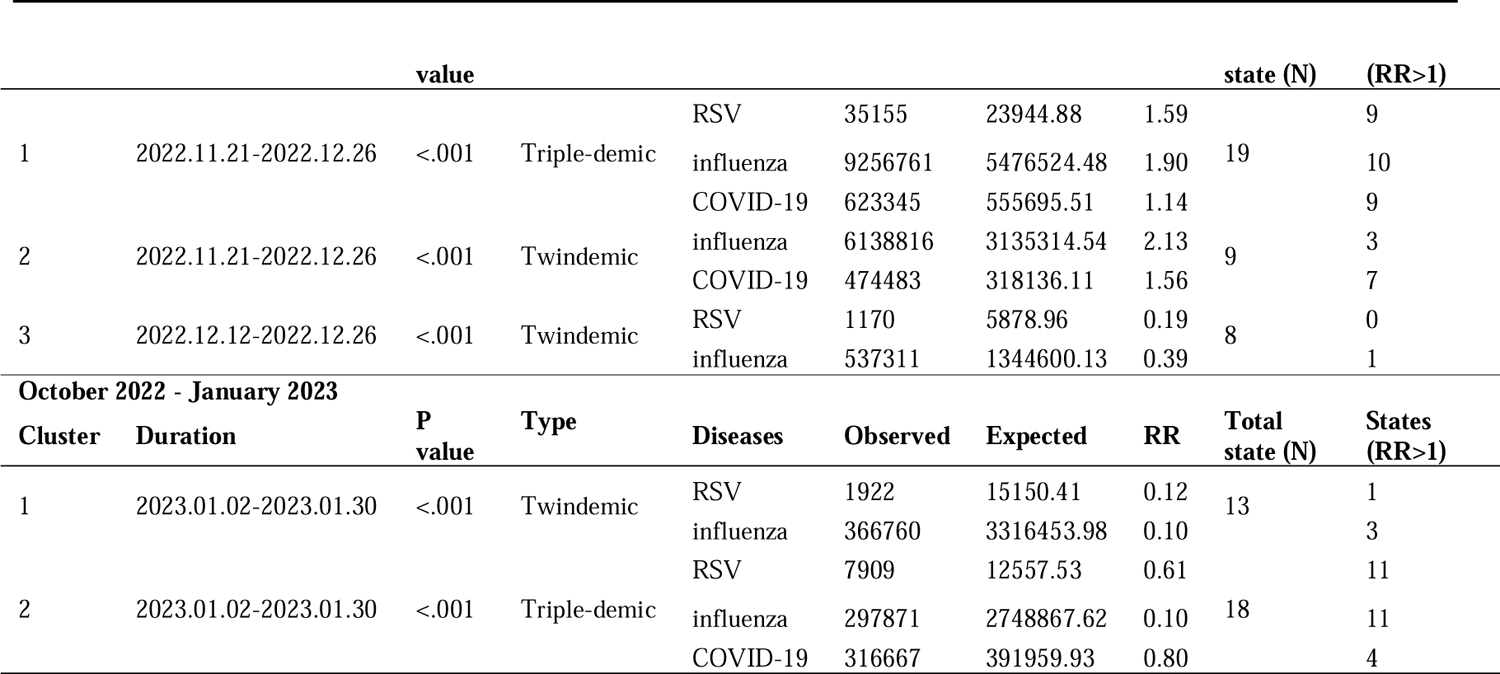
Space-time multivariate clusters from October 2022 to January 2023 (RR = relative risk).

### 3.3 Variations of States with Multiple-demic Risk

Table 4 illustrate the states with high RR (RR > 1) for both or all three diseases at various stages in winter of 2022. In October, 15 states in the eastern US demonstrated elevated risks of twindemic or triple-demic, with nearly half of them being categorized as states with twindemic risks of COVID-19 and RSV, while only 2 states, Kentucky and West Virginia, were classified as states with triple-demic risks. As of November, the number of states with twindemic or triple-demic risks increased to 18 in which 9 states exhibited twindemic risks of COVID-19 and RSV, 2 states (Minnesota and West Virginia) demonstrated high triple-demic risks, and 2 states (Mississippi and Washington) exhibited twindemic risks of flu and RSV. However, the number of states with twindemic risks of COVID-19 and flu increased from 3 to 5, primarily located in the eastern US (Figure A2). In December, the situation deteriorated for 22 states with twindemic or triple-demic risks. The numbers of states with risks of triple-demic increased from 2 before December to 4 at this stage, namely Minnesota, Nebraska, New Mexico, Ohio. The spatial pattern revealed that the twindemic or triple-demic risks spread from east to west, with an increasing number of western states experiencing the twindemic or triple-demic risks. Although 2 low-risk multivariate clusters emerged by the end of Jan 2023 (Figure 5d), a total of 17 states had twindemic and 4 states had high risks of triple-demic (i.e., New Mexico, North Dakota, Ohio) by the end of January (Figure A2d).

**Table 4.** Summary of States with Multiple-demic Risk between October 2022 and January 2023.

|  | October | November | December | January |
| --- | --- | --- | --- | --- |
| **Twindemic (COVID-19 & Flu)** | Delaware, North Carolina, Tennessee | Kentucky, New York, North Carolina, Pennsylvania, Tennessee | Kentucky, New Jersey, New York, Oklahoma, Pennsylvania, Tennessee | Alabama, Louisiana, New Jersey, New York, Oklahoma, Tennessee |
| **Twindemic (COVID-19 & RSV)** | Connecticut, Minnesota, New Mexico, Ohio, Pennsylvania, Vermont, Wisconsin | Colorado, Connecticut, Delaware, Hawaii, Massachusetts, New Mexico, North Dakota, Ohio, Wisconsin | California, Colorado, Connecticut, Hawaii, Massachusetts, North Dakota, Wisconsin | California, Connecticut, Hawaii, Massachusetts, Wisconsin |
| **Twindemic (Flu & RSV)** | Georgia, Mississippi, Texas | Mississippi, Washington | Iowa, Mississippi, Montana, Oregon, Washington | Iowa, Minnesota, Mississippi, Montana, Nebraska, Oregon, Washington |
| **Triple-demic** | Kentucky, West Virginia | Minnesota, West Virginia | Minnesota, Nebraska, New Mexico, Ohio | New Mexico, North Dakota, Ohio |

## 4 Discussion

### 4.1 Principal Findings

In this study, we applied univariate and multivariate STSS to retrospect the situation of COVID-19, influenza, and RSV from October 2021 to January 2022, and identify emerging clusters of “triple-demic” in the US from October 2022 to January 2023. To our knowledge, this is the first study to provide a near real-time approach to study the spatiotemporal variations of co-circulation of COVID-19, flu, and RSV in the US, thus providing evidence for improving the surveillance and control of the emerging “triple-demic” situation.

Generally, the situation of influenza and RSV became more severe during the winter of 2022, with significant increase of confirmed cases, while the number of COVID-19 cases maintained at a relatively low level during the winter of 2022 compared with the situation during the winter of 2021. By using retrospective space-time cluster analysis, we identified 3 high-risk clusters of COVID-19 during January 2022, a high-risk cluster of influenza from November to December 2021, and 2 high-risk cluster of RSV from October 2021 to January 2022. Moreover, a high-risk twindemic cluster of influenza and RSV were detected in the central US from November to December 2021, while certain eastern states experienced relatively low risk of influenza and RSV outbreak in the late winter of 2021.

In terms of the prospective STSS results during the winter of 2022, the univariate space-time cluster analysis revealed that high-risk clusters of COVID-19 and flu were primarily concentrated in the eastern states between October 2022 and December 2022. The western states underwent a transition from low risk in October 2022 to high risk in December 2022, and ultimately returned to low risk in January 2023. Throughout our study period, high-risk clusters of RSV were observed in various western and eastern states, with a particularly large cluster emerging in the western and central US during November. Notably, the southeastern regions experienced a sustained low risk of RSV throughout the winter of 2022, while many southwestern states transitioned to low risk of RSV starting in early January 2023, indicating a gradual improvement in the situation.

The multivariate prospective analyses revealed that the southern US experienced a high-risk cluster of RSV and influenza from October to November 2022. In late October 2022, a high-risk cluster of triple-demic emerged in the eastern US, which subsequently transitioned into a high-risk twindemic cluster of COVID-19 and influenza by late November 2022. This indicates that many eastern states, particularly those on the eastern coast, were at a high-risk of exposure to both COVID-19 and influenza between October and November 2022, highlighting the need for greater concern and attention. Furthermore, our analysis identified a large high-risk cluster of triple-demic that emerged in the central US from late November 2022. This suggests that 19 states were at a relatively high risk of experiencing triple-demic outbreaks. However, the situation of twindemic influenza and RSV in southeastern states began to improve from early December 2022. Additionally, the multiple-demic situation started to improve in many states from early January 2023. Noticeably, we found that the spatiotemporal patterns of multivariate clusters shared partial similarity with the univariate cluster of flu, which probably implies that flu overpowered the other two diseases. Throughout the winter of 2022, the number of states at high-risk for twindemic or triple-demic increased from October to the end of January, with a deteriorating trend from east to west. We suppose that the emerging outbreak of multiple-demic in the winter of 2022 was probably due to (1) a lack of recent immune responses to other virus after the public health interventions of COVID-19^39^; (2) inconsistent seasonal changes in meteorological factors which lead to increasing seasonal impacts of respiratory viruses^40^; (3) new strains of the pathogens which could evade previously established immune defenses^9^.

### 4.2 Strengths and Implications

The advantage of this study over prior works is that we provided a novel insight for spatiotemporal surveillance of the dynamics and characteristics of the “triple-demic” at the state level in the US. During the winter of 2022, the “triple-demic” also strained healthcare system in Canada^41^, UK^42^ and Singapore^43^. It is urgent to explore the extent of the impact of the three diseases and their modes of transmission. Our analysis can help monitor the “triple-demic” from spatiotemporal perspective, which can be also applied in other countries experiencing outbreaks of these three diseases. Additionally, our analysis provides insights for public health authorities to prioritize resource allocation and mitigate potential outbreaks of multiple epidemics worldwide. Since the concept of a “triple-demic” is a relatively new one and has not yet been experienced on a global scale, few studies have specifically examined the topic. Prior studies generally paid more attention to investigating the outbreak pattern of two out of three respiratory infections simultaneously^44–46^. For instance, during the COVID-19 pandemic in Australia, there were unusual and large outbreaks of RSV that persisted from spring to summer^47^.

Previous studies have generally found that the co-occurrence of multiple infectious diseases can have a compounding effect on public health outcomes, including increased illness, hospitalizations, and deaths^48, 49^. Consistent with these findings, our study detected several states and clusters with high risks of two or three diseases, especially in the eastern US. This highlights the potential for an increased medical burden in these areas and guarantees greater attention. Hence, we should remain vigilant about the potential exacerbation of these three diseases. Our methods can also be used for further real-time monitoring to promptly detect the possible emergence of multiple-demic clusters. Our findings suggest that states that suffered from multiple-demic should prioritize the distribution of influenza vaccines. A viable suggestion at this stage is to prioritize the widespread distribution of vaccines and fully prepare the medical reserves to prevent overwhelming medical institutions. When vaccination rates have increased significantly, further adjustments to interventions can be considered to transition back to normal life^50^. These policy implications may be applicable to other countries.

### 4.3 Limitations

There were several limitations in our study. First, RSV cases are continuously updated by the CDC, and timely data availability from each state is not always possible due to a shortage of labs and tests. Second, under-reporting and potential misdiagnosis due to clinical symptom similarities among the three viruses remain a concern^51, 52^. Third, we did not consider sociodemographic factors that play a crucial role in disease transmission^53^. For example, RSV-related deaths are most common in children and the elderly in the winter of 2022^54, 55^. Incorporating sociodemographic factors (e.g., adjusted rates based on socioeconomic status) in the space-time scan statistic might generate more accurate RR estimates. Moreover, to enhance the identification of potential multiple-demic situations and understand the underlying mechanisms contributing to spatiotemporal disparities in multi-demic risks, high spatial resolution data (i.e., county) is recommended for future study to improve usability and accuracy.

## 5 Conclusion

In conclusion, our study shed light on the processes that contributed to the “triple-demic” in the US states over space and time. Our approach can be utilized for continuous surveillance the “triple-demic” dynamics with the latest data, and to facilitate timely adjustments of domestic and interregional public health interventions that aimed at preventing further deterioration of the multiple-demic situation.

## Contributor

WL, QL, YZ, SL, and ZL developed the study protocol, and designed the study. QL, YZ, and YR wrote the first draft. QL obtained the data and conducted the software. YZ and WH visualized the results. WL, SL, SP, and ZL reviewed and commented on the paper. WL and SL supervised the work. All authors revised the manuscript and approved the final paper.

## Data Availability

All data produced in the present work are contained in the manuscript

## Appendix

### Spatiotemporal patterns of space time clusters in February 2023

**Table A1.** Space-time univariate clusters of COVID-19, influenza, and RSV from October 2022 to February 2023 (RR = relative risk).

| Cluster | Duration | P value | Diseases | Observed | Expected | RR | Total state (N) | States (RR>1) |
| --- | --- | --- | --- | --- | --- | --- | --- | --- |
| 1 | 2023.01.30-2023.02.27 | <.001 | COVID-19 | 5170 | 104695.02 | 0.049 | 1 | 0 |
| 2 | 2022.12.19-2023.02.27 | <.001 | COVID-19 | 1344668 | 1011763.56 | 1.41 | 14 | 12 |
| 3 | 2023.01.23-2023.02.27 | <.001 | COVID-19 | 310983 | 483025.72 | 0.63 | 16 | 3 |
| 4 | 2022.12.26-2023.02.27 | <.001 | COVID-19 | 230912 | 148356.35 | 1.58 | 4 | 3 |
| 1 | 2023.01.02-2023.02.27 | <.001 | influenza | 448116 | 4935213.88 | 0.081 | 13 | 3 |
| 2 | 2023.01.09-2023.02.27 | <.001 | influenza | 404088 | 4545098.70 | 0.080 | 18 | 11 |
| 1 | 2022.12.19-2023.02.27 | <.001 | RSV | 3213 | 27901.60 | 0.10 | 15 | 1 |
| 2 | 2023.01.09-2023.02.27 | <.001 | RSV | 5937 | 20735.85 | 0.26 | 10 | 3 |

**Table A2.**
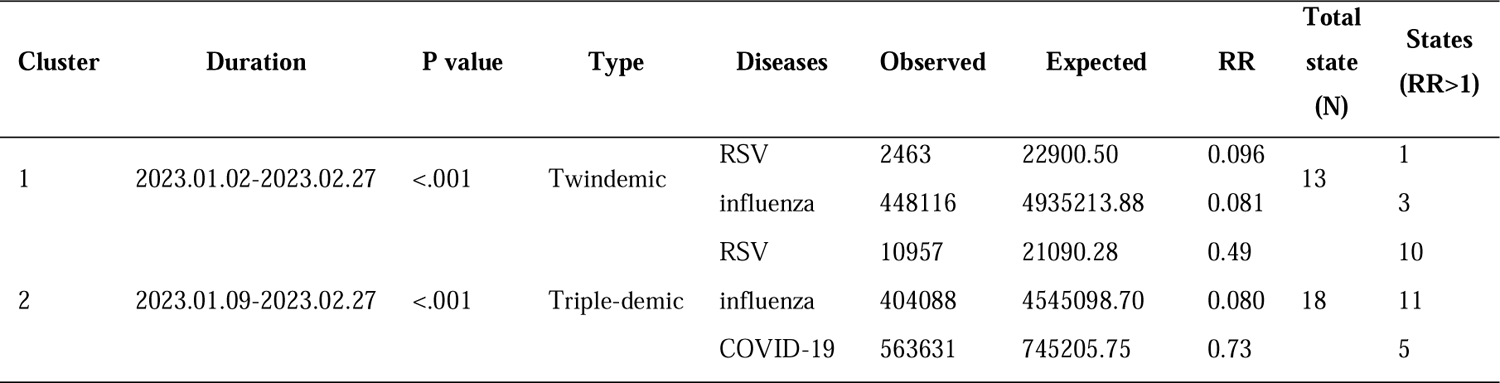
Space-time multivariate clusters from October 2022 to February 2023 (RR = relative risk).

**Table A3.**
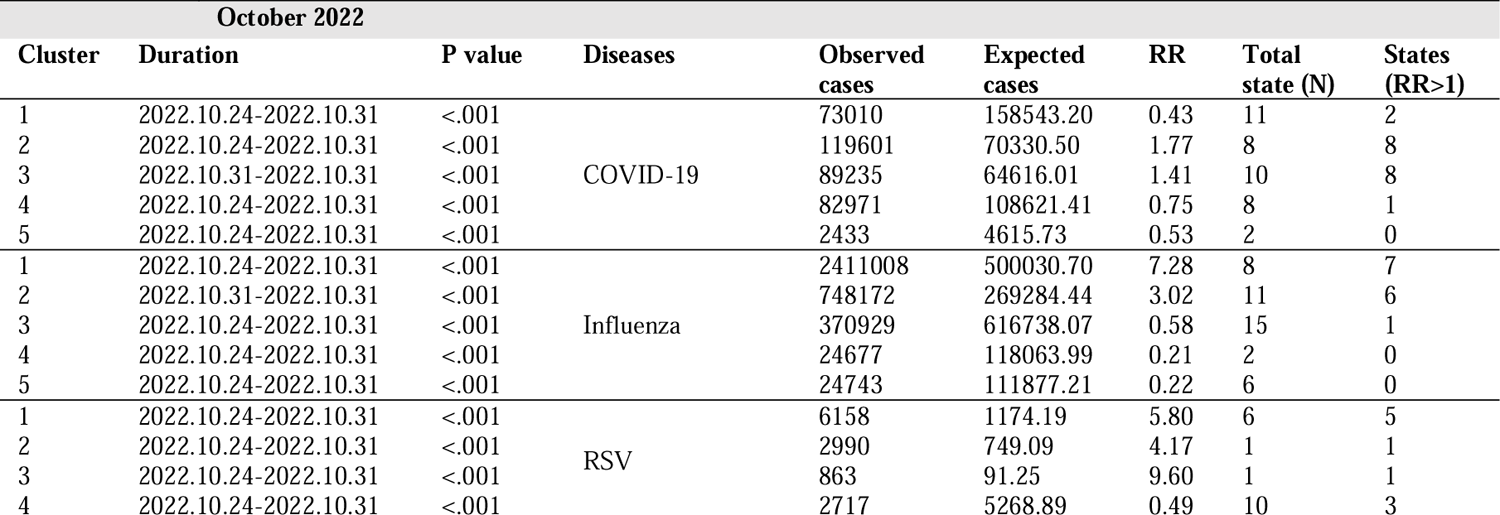
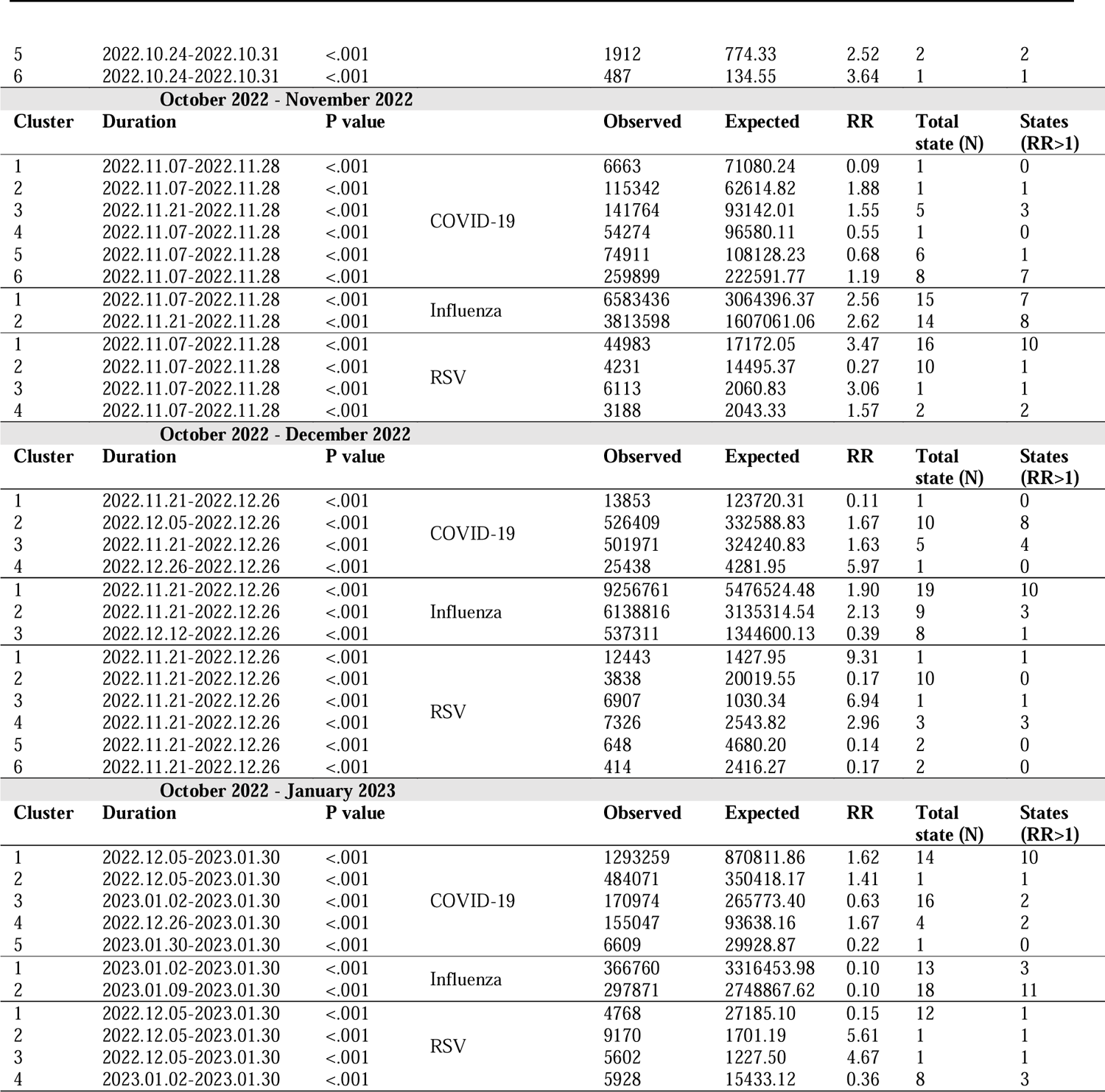
Space-time univariate clusters from October 2022 to January 2023 (RR = relative risk).

**Figure A1.**
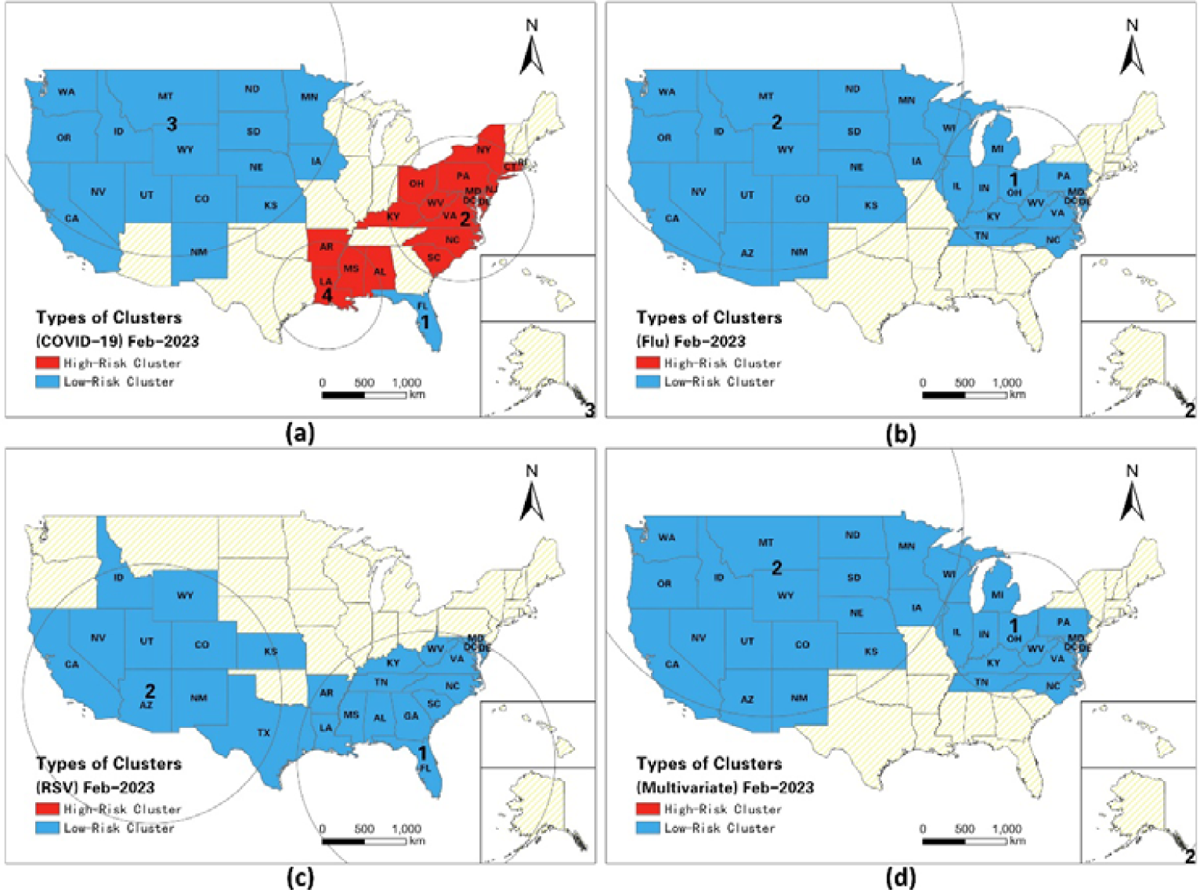
Spatial Patterns of space-time clusters in February 2023

**Figure A2.**
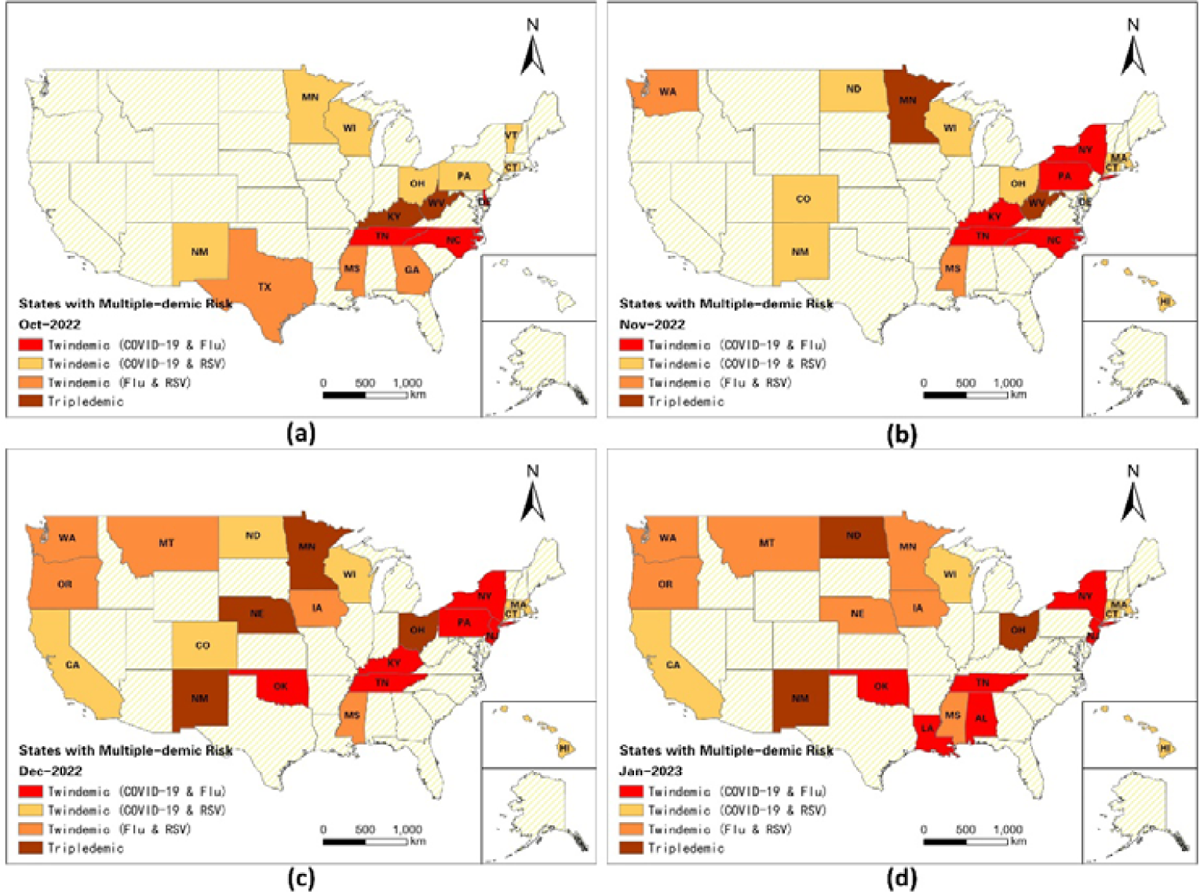
Spatiotemporal distribution of states with multiple-demic risks during the winter of 2022.

